## supplemental figure for "Kangaroo Mother Care prior to clinical stabilisation: Implementation barriers and facilitators reported by caregivers and health care providers in Uganda"

**Figure 1. Conceptual framework for schematic analysis of barrier & facilitators of KMC, by six adapted health system building blocks.**

**Family & community support & involvement**

- Family member present in the hospital
- Cultural beliefs & practices

**Human resources for Health**

- Staffing levels & competency
- Ability to support caregivers.

**Medical supplies & devices**

- Availability of medicines & devices
- Timely treatment

**Infrastructure and design**

- KMC space including in high dependency area, maternal beds, and privacy.
- Provision of washrooms for families

**Financing**

- Adequate financing to enable high coverage and quality of care.

**Health facility leadership**

- Support for staffing & training
- Overall enabling policy environment, with accountability
